# Excessive Corneal Endothelial Single Cell Loss Following Endothelial Injuries

**DOI:** 10.1101/2024.09.09.24313319

**Authors:** Yuan Kai Fu, Matthew Lin, Kuo-Hsuan Hung, Lung-Kun Yeh, Hsin-Yuan Tan

**Affiliations:** Department of Ophthalmology, Chang Gung Memorial Hospital, Linko Branch, Taiwan; Taipei American School, Taipei, Taiwan; College of Medicine, Chang Gung University, Taoyuan, Taiwan

**Author notes:** **Correspondence for reprints:** Hsin-Yuan Tan, MD, PhD, Associate Professor, Department of Ophthalmology, Chang Gung Memorial Hospital, Linko, Taiwan, No. 5, Fu-Shin St. Kwuei-Shang Shiang, Taoyuan City 333, Taiwan.

**Keywords:** corneal endothelium, endothelial cell loss, cataract surgery, corneal trauma, corneal endothelial dystrophy

## Abstract

Corneal endothelial dysfunction is the main cause for more than 50% of corneal transplantations. Human corneal endothelial cells are generally viewed as non-proliferative *in vivo*. Any injury that results in endothelial loss exceeding the critical threshold can cause irreversible endothelial functional decompensation, leading to corneal edema and vision loss. Currently, the mainstay treatment for irreversible corneal dysfunction is corneal transplantation. In this work, using well-established imaging technique of specular microscopy, we revisited the endothelial damage following three common corneal endothelial injury scenarios: post-cataract surgery, endothelial dystrophy, and corneal penetrating injury. We identified unexpected, stochastic single-cell loss in the corneal endothelium following primary injuries, persisting well beyond the expected wound healing period, a phenomenon that has not been previously highlighted. This finding offers a potential explanation for the chronic endothelial cell loss following a primary injury. Further investigation could provide valuable insights for improving clinical management strategies for corneal endothelial dysfunction.

## Introduction

Corneal endothelial dysfunction is one of the leading causes of corneal transplantation, underscored by its prevalence in over 50% of such cases[1]. The corneal endothelium is a single-layered cell lining the inner side of the cornea, which responds to corneal nutrition and the regulation of corneal hydration for keeping the cornea in a relatively dehydrated status, for maintaining the cornea in a transparent status[2]. Unlike corneal epithelium, corneal endothelium does not replicate *in vivo* at normal physiological status, even though it possesses the capability of proliferation[2]. Under normal physiological conditions, the adult human corneal endothelial cell density is approximately 2500-3000 cells/mm^2^[3], and decreases 0.6% annually by age[4]. Any injury or disease that causes additional endothelial cell damage can lead to an irreversible decompensation of the cornea. When the endothelial cell density drops below a critical threshold of approximately 500 cells/mm^2^[5], it results in the swelling of the cornea, loss of corneal transparency, and consequential vision loss. The corneal endothelium’s non-proliferative nature and limited regenerative capacity cause a significant challenge for endothelial regeneration. Although some pharmaceutical approaches have been developed recently, currently, the mainstay for treating corneal endothelial decompensation is still allogeneic transplantation[6].

The comprehensive mechanism of corneal endothelium wound healing remains unclear[2]. Unlike corneal epithelium, the endothelial cells are mostly arrested in the G1 phase, and heal wounds by cell enlargement and migration of adjacent cells, but not by proliferation [7]. Therefore it is believed that when the damaged area is too large to be completely covered, and the residual cell density drops beyond the threshold to maintain normal watering pumping function. The cornea then decompensates, becoming swollen and not transparent, leading to the loss of vision. Pseudophakic bullous keratopathy (PBK), Fuchs’ endothelial dystrophy (FED), and trauma are the common clinically-encountered endothelial diseases that cause irreversible endothelial decompensation[1]. PBK refers to irreversible corneal endothelial dysfunction following cataract extraction and intraocular lens implantation surgery [8]. The prevalence of PBK is approximately 1% [9]. The primary cause of endothelial cell loss in PBK is surgical trauma, which results from the thermal damage induced by the increased local temperature associated with the phacoemulsification probe, and the turbulent flow, the ultrasound energy, and the production of free radical and oxidative stress during the procedure of phacoemulsification of lens[8]. FED is the most common form of endothelial dystrophy which is characterized by the endothelial cell loss and development of guttae, the excrescences of Descemet’s membrane. FED progresses slowly within time and will expand to significant endothelial cell loss, subsequential loss of corneal deturgescence, and irreversible corneal edema [10]. The prevalence of FED varies greatly, with an estimated 7.33% globally[11]. In some clinical scenarios of corneal endothelial injury, we did observe that the corneal endothelial cell loss progresses even if the causative incident did not proceed. The onset of bullous keratopathy following cataract surgery and Argon laser iridotomy was reported at mean 148.4 and 107 months respectively[12]. The exact mechanism for the progression of endothelial cell loss beyond normal aging loss following primary insult remains uncertain. Therefore in this observational study, we revisit the endothelial cell loss in three common causes of corneal endothelial injury, including post cataract surgery, endothelial dystrophy, and penetrating corneal injury, to identify the potential insights into corneal endothelial wound healing.

## Materials and Methods

*Study Design* This cross-sectional observational study received approval from the institute review board of Chang Gung Memorial Hospital, Linko, Taiwan. We included patients with three common causes of endothelial injury, including post-cataract surgery, corneal endothelial dystrophies, and penetrating corneal injury. Basic data on patient demographics, diagnosis, and treatment history were reviewed and collected. Subjects diagnosed with active corneal diseases other than endothelial dystrophies, as well as those who had undergone corneal transplantation were excluded. The patients with severe corneal edema observed under slit-lamp biomicroscopy that confounded specular microscopic examinations, were also excluded. The non-contact *in vivo* specular microscopy (CEM-530, Nidek, Gamagori, Japan) was employed for endothelial cell morphological analysis. The endothelial photographs were captured at five distinct regions of the cornea for each patient using a standard magnification of 400X (**Fig.1**). These regions included the central point - with the fixation light aimed directly at the center - and four paracentral points (upper, lower, nasal, and temporal). For the paracentral points, the fixation light targeted a paracentral zone with a diameter of 1.3mm at specific angular positions. In the right eye (OD), these positions were at 90°, 270°, 0°, and 180°, while in the left eye (OS), they were at 90°, 270°, 180°, and 0°, respectively. The obtained images were processed using the manufacturer-provided software, which calculated morphometric parameters including cell density (ECD), coefficient of variation (CV), and percentage of hexagonal cells (HEX) through the inbuilt image analysis system.

**Fig 1.**
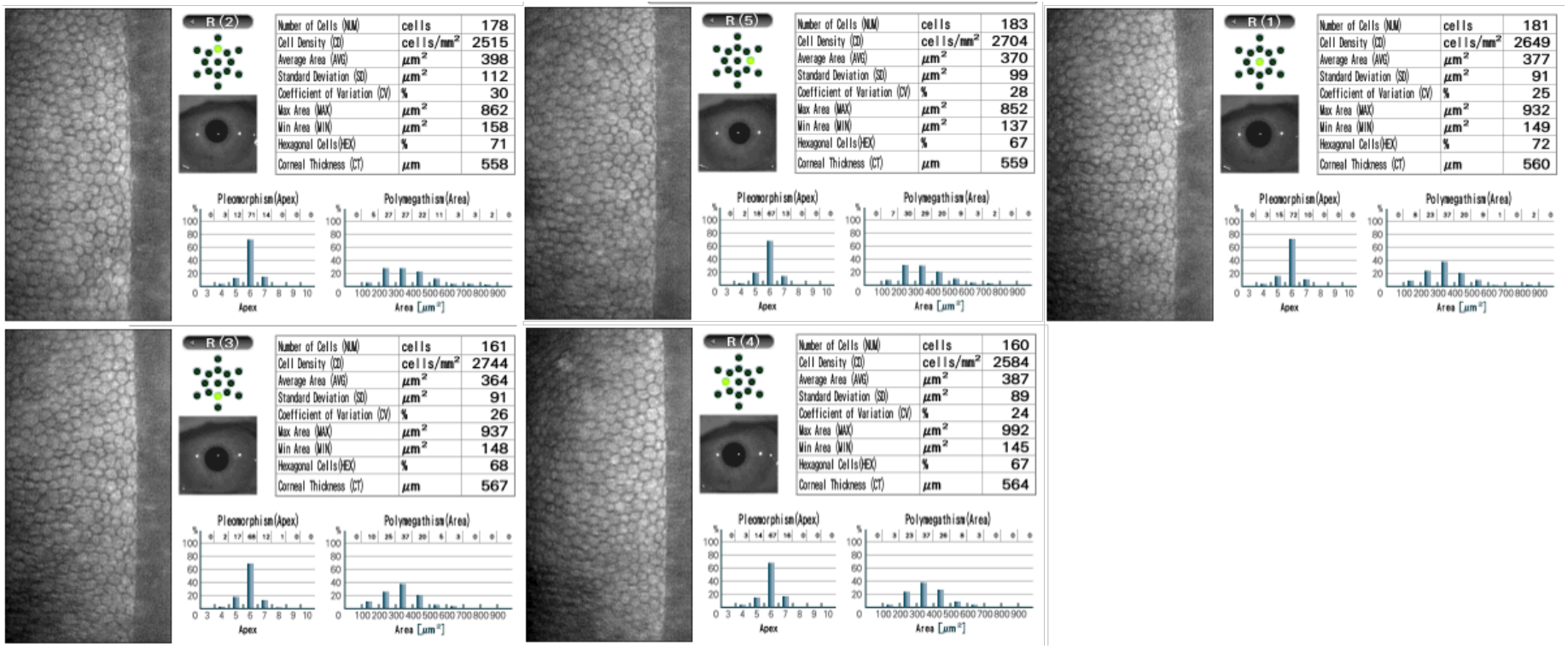
Representative corneal endothelial images captured at five distinct locations: the central point and four paracentral points at 0^°^, 90^°^, 180^°^, and 270^°^ (magnification 400X).

*Statistical Analysis* All statistical analyses in this study were conducted using SPSS version 29.0 (SPSS Inc., Chicago, IL, USA). Descriptive statistics were applied to the basic data, including age, sex, and laterality of the eye across the three groups: post-cataract surgery, endothelial dystrophy, and trauma. The endothelial morphometric parameters included ECD, CV, and HEX from 5 distinct regions for each case, were also compared between the injured eye and the non-injured fellow eye, using the Wilcoxon Signed Ranks Test , two-sided.

## Results

In this cross-sectional observational study, we enrolled patients with three common causes of endothelial injury, including post-cataract surgery, corneal endothelial dystrophies, and penetrating corneal injury. The endothelial photographs taken by specular microscopy with unclear images, where endothelial morphology could not be detected or endothelial parameters could not be analyzed, were excluded from the study. We included 83 patients in this observational study. The mean age was 63.2 years-old, with 52 females and 31 males. We collected effective endothelial data from 32 eyes who had undergone cataract surgery without having endothelial dystrophy , 32 eyes with endothelial dystrophy who had not undergone cataract surgery, 14 eyes with endothelial dystrophy and had received cataract surgery, and 7 eyes with penetrating corneal injuries. The mean duration from primary injury to examination was 26.57 ± 55.69 weeks, ranging from 1 day to 249.29 weeks for cataract surgery cases, and 28.07 ± 29.90 weeks, ranging from 1 day to 77.57 weeks for trauma cases. The mean endothelial cell density at the center was 2,138.91 ± 869.34 cells/mm^2^ in the post-cataract surgery group (n=46), 1,999.48 ± 763.91 cells/mm^2^ in the FED group (n=46), and 1,854.86 ± 551.85 cells/mm^2^ in the trauma group (n=7). The endothelial morphometric parameters are listed in **Table 1**. We compared the eyes post-cataract surgery with their uninjured fellow eyes and found no significant differences in most parameters of endothelium after injury at five distinct regions of cornea (**Table 2**). A significant increase in CV value was observed in the upper and temporal regions of post-cataract surgery eyes (p<0.05), although it remained within the normal range. Due to the small sample size of 7 trauma eyes and the large variation within the group, we did not perform statistical comparisons. Unexpectedly, we observed a distinct pattern of stochastic single-cell loss in the injured corneas (21.18%) in all three endothelial injury scenarios, even after a duration of two years after the primary injury (**Fig. 2**). In contrast, no single-cell loss was observed in the uninjured fellow eyes of cataract surgery and trauma cases. Although the selected regions may not fully represent the response of the entire endothelium to injury, we nonetheless highlighted a phenomenon of stochastic single-cell loss occurring after the primary injury, even well beyond the expected wound healing period.

**Table 1.** Selected endothelial parameters from five distinct corneal regions following three common endothelial injuries.

| Injury Type | Location | N | ECD (cells/mm<sup>2</sup>) | CV(%) | HEX(%) | Duration(weeks) |
| --- | --- | --- | --- | --- | --- | --- |
| Endothelial dystrophy | Central | 46 | 1999.48±763.91 | 36.74±14.45 | 56.28±22.36 | N/A |
|  | Superior | 29 | 2157.31±682.3 | 33.10±8.32 | 60.62±10.08 | N/A |
|  | Inferior | 30 | 2445.03±519.45 | 34.3±8.78 | 61.67±15.67 | N/A |
|  | Nasal | 30 | 2366.43±534.42 | 32.53±7.67 | 62.8±12.61 | N/A |
|  | Temporal | 27 | 2033.93±766.8 | 34.85±11.75 | 56.85±21.25 | N/A |
| Post-cataract surgery | Central | 46 | 2138.91 ±869.34 | 34.98±14.42 | 58.63±18.20 | 186.46±389.85 |
|  | Superior | 41 | 2192.12±866.3 | 33.27±8.40 | 58.22±15.36 | 184.59±414.20 |
|  | Inferior | 42 | 2395.52±1001.5 | 32.02±12.42 | 58.67±15.19 | 186.03±408.27 |
|  | Nasal | 43 | 2384.53±704.05 | 32.42±8.36 | 62.23±10.38 | 181.22±403.42 |
|  | Temporal | 42 | 2205.12±798.02 | 35.69±12.02 | 62.60±14.50 | 185.69±408.41 |
| Corneal penetrating injury | Central | 7 | 1854.86 ±551.85 | 27.57±4.20 | 61.71±27.73 | 196.5±209.29 |
|  | Superior | 5 | 2557.0±273.57 | 29.0±3.0 | 67.8±7.05 | 113.6±153.68 |
|  | Inferior | 3 | 2484.33±167.31 | 28.67±7.09 | 67.33±7.23 | 159.33±198.48 |
|  | Nasal | 4 | 2526.5±267.13 | 29.5±6.24 | 66.25±3.95 | 130.75±171.85 |
|  | Temporal | 5 | 1801.0±707.57 | 36.2±11.41 | 46.8±27.37 | 97.2±164.12 |

Duration: duration from primary injury to examination; ECD: endothelial cell count; CV: coefficient of variant; HEX: hexagonality; N: number of photographs

**Table 2.** Comparisons of selected endothelial parameters from five distinct corneal regions following cataract surgery and in the fellow eyes.

|  | Non-operated eye | Operated eye | p | Non-operated eye | Operated eye | p | Non-operated eye | Operated eye | p | Non-operated eye | Operated eye | p | Non-operated eye | Operated eye | p |
| --- | --- | --- | --- | --- | --- | --- | --- | --- | --- | --- | --- | --- | --- | --- | --- |
| Location | Central |  |  | Superior |  |  | Inferior |  |  | Nasal |  |  | Temporal |  |  |
| **n** | 24 |  |  | 22 |  |  | 21 |  |  | 22 |  |  | 22 |  |  |
| **Duration**<br>(weeks) | 53.09±80.68 |  |  | 39.29±36.37 |  |  | 39.6±37.50 |  |  | 39±36.64 |  |  | 38.48±36.91 |  |  |
| **ECD**<br>(cells/mm<sup>2</sup>) | 2506.63<br>±345.12 | 2473.5<br>±455.82 | 0.152 | 2500.77<br>±274.87 | 2514.50<br>±368.08 | 1 | 2607.05<br>±312.61 | 2603.52<br>±561.53 | 0.664 | 2589.18<br>±313.46 | 2593.14<br>±371.50 | 0.548 | 2432.09<br>±498.70 | 2463.09<br>±396.34 | 0.57 |
| **CV**(%) | 30.0± 6.50 | 30.46± 8.53 | 0.286 | 29.55±6.01 | 33.36±6.86 | 0.019 | 30.05±4.34 | 32.10±6.27 | 0.167 | 29.09± 5.62 | 31.91± 6.63 | 0.081 | 28.95±4.60 | 33.95± 8.72 | <.001 |
| **HEX**(%) | 66.5±8.76 | 65.63±15.73 | 0.839 | 68.5±7.04 | 64.05±8.85 | 0.078 | 65.57±6.45 | 63.90±9.21 | 1 | 67.32± 6.90 | 64.86± 8.65 | 0.413 | 67.36±5.66 | 67.59±7.02 | 0.702 |

P value by Wilcoxon Signed Ranks Test between operated group and non-operate fellow group

**Fig 2.**
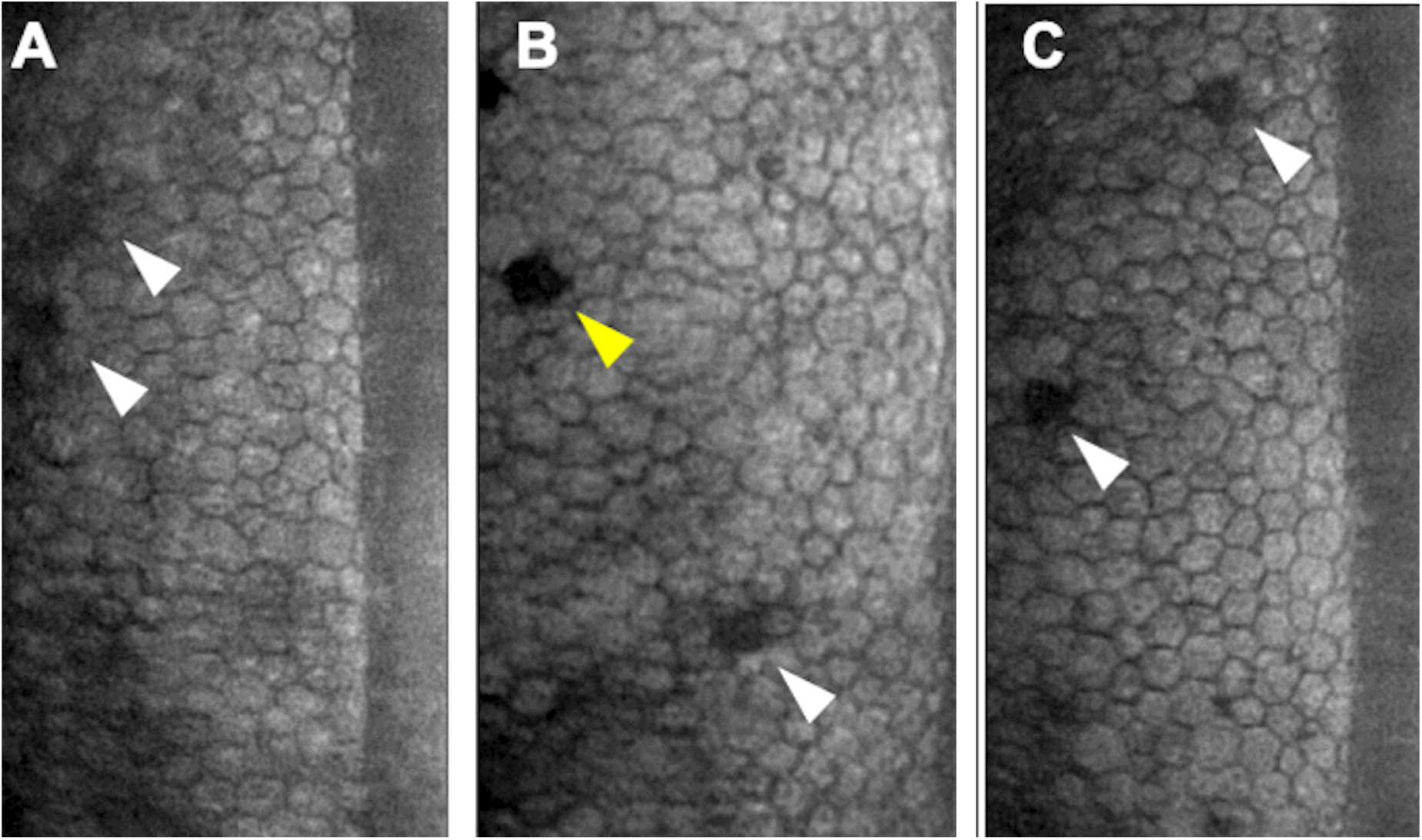
Representative endothelial photographs captured by specular microscopy, showing single-cell loss observed across all three injury scenarios. **(A)** Post-cataract surgery, two years after surgery; **(B)** Fuchs’ endothelial dystrophy; **(C)** Penetrating corneal injury, five weeks post-injury. (White arrows indicate single endothelial cell loss; yellow arrow indicates guttae)

## Discussion

In our observational study, we investigated endothelial morphology following three common endothelial injuries: posterior-cataract surgery, FED, and trauma. We discovered an interesting new phenomenon, where excessive, random single-cell loss was consistently observed in injured corneas across different types of endothelial injuries (21.18%), which was absent in normal fellow eyes. Apart from corneal endothelial dystrophy, although the primary etiological factors differ, a common observation is the persisting disease progression that exceeds the normal aging changes, even after the main pathological process ceases. The exact mechanism of corneal endothelium wound healing is still unclarified [13]. It remains unclear how a primary endothelial injury leads to a chronic progression of endothelial cell loss, eventually resulting in irreversible endothelial decompensation requiring transplantation. The chronic endothelial cell loss following routine cataract surgery is an example. Chronic progressive endothelial cell loss following cataract surgery has been widely reported [14-16]. The majority of studies evaluate the endothelium within 1 year postoperatively, with varying reported rates (2.3-18.19%)[14, 17, 18]. The preoperative patient condition and the surgical techniques all contribute to the large variation [14, 17]. Despite the modern refinement of cataract surgery, a 2.06% endothelial cell loss rate per year[15] following uncomplicated cataract surgery, which significantly outraces physiological aging (0.6%)[19], has still been reported in a long-term (10 years) follow-up study. And another 7-year prospective study reported a total 23.47% endothelial cell loss, with an affirmed progression over time [14]. The endothelial cell loss following cataract surgery has been proposed in two stages. The immediate post-operative endothelial cell loss is believed to be directly related to surgical procedures[20]: thermal and mechanical damages caused by phacoemulsification, contact with lens fragments during surgery, and inflammation are the main reasons for immediate endothelial cell loss following cataract surgery[21-24]. After the initial endothelial wounding is morphometrically stabilized 3 months following cataract surgery [25], the continued and accelerated long-term endothelial cell loss has been hypothesized to be related to subclinical inflammation, decreased innervation, loss of vitreous, and possibly a tendency for endothelial remodeling — that has not been confirmed until recently [15, 16]. In the 10-year follow-up study, the nuclear firmness and early postoperative corneal edema are proposed as predictive factors for long term endothelial cell loss[15]. However, their findings are to an extent contradicted with the late endothelial remodeling hypothesis, which states there is no difference in the morphological indices between pre- and post-operative endothelium[15]. Moreover, in the 7-year follow-up study, Lundberg confirmed the trend of chronic endothelial cell loss[14]. Interestingly, the late endothelial cell loss is less in corneas with significant immediate postoperative corneal edema, and the total cell loss is similar regardless of the severity of early corneal edema. It therefore implies a possibility that the primary endothelial injury per se initiates the long-term endothelial cell loss, regardless of the severity of early corneal edema [14]. Another clinical example of significant chronic corneal endothelial loss is the Argon laser iridotomy (ALI) induced bullous keratopathy [26]. The ALI is a well-established procedure for treating and preventing angle-closure glaucoma [27]. The approximated incidence of ALI-induced corneal edema was estimated at 1.8%[26]. It has been reported as high as 20% in etiology for corneal transplantation in Japan [26]. The mean duration between ALI and the first visit due to corneal edema was 5.5-7.4 years [26, 28]. Several hypotheses have been proposed regarding the pathogenesis of ALI-induced cornea. The direct focal laser damage of endothelium, the increase of aqueous humor temperature, high laser energy, breakdown of the blood-aqueous barrier, the inflammation, iris pigment dispersion, and the shear stress caused by the turbulent aqueous stream affecting the endothelium [26, 29, 30] are all the causative factors contributing to endothelial damage. A case of focal corneal decompensation remote from ALI site has also been reported [31]; it was speculated to be related to ALI-generated debris accumulation far from ALI site.

However, an unidentified chronic damaging mechanism is still likely the main cause of corneal edema as it manifests mostly as a late-onset issue. In this observational study, we revisited the endothelial morphometric changes following three common endothelial injuries. We observed an interesting new phenomenon of stochastic single-cell loss in the injured cornea (21.18%), consistently across all three common clinical endothelial injury scenarios, which was not seen in the normal fellow eyes. The cell loss occurred at regions distant from the main lesion **(Fig. 2A)** as well as near the guttae **(Fig. 2B)**. It could be observed as late as two years following primary injury insult. To our knowledge, it is the first report that highlighted this morphological finding of the endothelium following primary endothelial injuries. We hypothesize that the primary endothelial wound healing process might somehow induce secondary cell loss outside the primary wound during the healing process. Stochastic loss of healthy, possibly migrating, endothelial cells can occur during this process. And the secondary single-cell loss can become another new wound, triggering further cascades of endothelial injury. When the generally accepted mechanisms of endothelial wound healing, including migration and enlargement of neighboring cells, and possibly, the induction of limited proliferative capacity of endothelium, all together fail to compensate the primary and secondary cell loss, an accelerated, chronic cell loss occurs, eventually leading to decompensation of endothelium. Further study will be conducted to clarify the mechanism for excessive stochastic cell loss following primary endothelial injury. The major limitation of our study is the lack of longitudinal follow-up data, as this is only an observational study. Additionally, although we captured images from five different regions of the endothelium, it will still be beneficial to have data covering a larger area to more comprehensively observe the wound healing response across the entire endothelium. In conclusion, our study revealed a novel phenomenon of stochastic single endothelial cell loss following common primary endothelial injuries, which continues well beyond the expected healing period. This finding offers insights into the unexplained chronic endothelial cell loss following an initial injury. Future research will aim to investigate the underlying mechanisms and potentially offer valuable information to improve the treatment strategies for corneal endothelial dysfunction.

## Data Availability

All data produced in the present study are available upon reasonable request to the authors

## Notes

### Competing Interest Statement

The authors have declared no competing interest.

### Funding Statement

This study did not receive any funding

### Author Declarations

IRB of Chang Gung Medical Foundation gave ethical approval for this work

